# Entitled to Property: Inheritance Laws, Female Bargaining Power, and Child Health in India

**DOI:** 10.1101/2021.06.20.21259224

**Authors:** Shahadath Hossain, Plamen Nikolov

**Author notes:** Contact information: Plamen Nikolov  and Shahadath Hossain.

## Abstract

Child height is a significant predictor of human capital and economic status throughout adulthood. Moreover, non-unitary household models of family behavior posit that an increase in women’s bargaining power can influence child health. We study the effects of an inheritance policy change, the Hindu Succession Act (HSA), which conferred enhanced inheritance rights to unmarried women in rural India, on child height. We find robust evidence that the HSA improved the height and weight of children. In addition, we find evidence consistent with a channel that the policy improved the women’s intrahousehold bargaining power within the household, leading to improved parental investments for children. These study findings are also compatible with the notion that children do better when their mothers control a more significant fraction of the family. Therefore, policies that empower women can have additional positive spillovers for children’s human capital. (*JEL* D13, I12, I13, J13, J16, J18, K13, O12, O15, Z12, Z13)

## I. Introduction

Height in early childhood is strongly predictive of cognitive ability (Case and Paxson, 2008), educational attainment (Currie, 2009), labor market outcomes (Smith, 2009; Persico et al., 2004), and occupational grade in later life (Case et al., 2009). Stunting, a key marker of severely impaired growth and low height-for-age, affects about 25 percent of children under five years of age in low- and middle-income countries (LMICs) (UNICEF, 2015). The prevalence of stunting and malnutrition is particularly acute in India, where 24 percent of the world’s stunted children live (WHO, 2021). India’s stunting rate stands at 31 percent, which stands as an outlier even when compared to other developing countries.

Although numerous factors – such as genetics, biology, and disease environment^1^ –influence height in early life, parents and resource allocation at the household level can play a crucial role (Rosenzweig and Schultz, 1982; Jayachandran and Pande, 2017). A key aspect of parental resource allocations relates to the locus of control in the family and decision-making power, especially women’s bargaining power (Thomas, 1990). Women spend more on nutrition (Imai et al., 2014), medical care (Maitra, 2014), childcare (Behrman, 1988; Strauss et al., 2000), and exhibit “maternal altruism” (Mason, 1986). Therefore, better female bargaining power at the household level can improve girls’ survival rates (Qian, 2008) and results in better anthropometric measures (Duflo, 2003).

In this paper, we explore the effect of the passage of an inheritance law, which confers improved inheritance rights to unmarried women in rural areas of India, on child height. In India, a predominantly rural country, land ownership is a critical determinant of economic and social status (Agarwal, 1994), and inheritance is the primary way to acquire it. However, conventional male-biased inheritance practices and cultural conservatism (Agarwal, 1994) engender limited earning opportunities and bargaining power for married women in India. Because of the potential to transform women’s inheritance rights, some Indian states amended the male-favored Hindu Succession Act (from now on HSA) by providing married women an equal share in ancestral property. Improved female bargaining power could improve human capital outcomes and gender inequality. Thus, enhanced inheritance rights for women may improve human capital outcomes and improved the status of daughters. It is this possibility that we explore in this paper.

Our estimation approach relies on the staggered adoption of the HSA amendment (hereafter, “HSAA”) across different states in India since 1956. We rely on two primary sources of variation related to a woman’s exposure to the HSAA. The first source of variation is the timing of a woman’s marriage. The reform in each state only affects unmarried women; thus, women who were married before the HSAA are unaffected by the policy.^2^ The second source of variation stems from the HSAA implementation, as some states amended the HSA first relative to those states that never amended the law before the 2005 federal amendment. The four treated states in our sample are Andhra Pradesh, Tamil Nadu, Karnataka, and Maharashtra. To answer the research question, we use data from the Indian National Family and Health Survey (NFHS), a cross-sectional survey with three rounds of data: Wave 1 (1992-1993), Wave 2 (1998-1999), and Wave 3 (2005-2006).

We find clear evidence that the passage of the HSAA improved, and substantially so, child health. Specifically, we find that the HSA reform improved child height, or the height-for-age (HFA), by 0.183 standard deviations. Similarly, we detect a substantial effect on weight, proxied by the WFA measure, by 0.276 standard deviations. Furthermore, and to shed additional light on where these improvements come from, we examine the impact on parental investments, and we find the HSA improved prenatal and postnatal investments. In terms of heterogeneous effects, we find evidence that the HSA positively affects the daughters’ health but only on short-term health outcomes, such as weight. We find no evidence that the HSA improved their height or parental care. In terms of the reform’s effects on higher birth order children, we find the HSA has no positive impact for them and may even have an adverse effect, via a reduction in the parental allocations, on the health of higher birth-order children. Finally, we find no evidence that HSA reform had any favorable impact on daughters of higher birth order.

Although we find strong evidence that the HSA improves child health, our findings show that these health benefits accrue mainly to first-born sons only. In India, preference for a first-born son can influence household resource allocations among children, especially in families of larger size.^3^ Existing research also documents that later-born children, and especially daughters, benefit less from parental investments. Familiar having more children could adversely affect each child’s health due to resource constraints (Booth and Kee, 2005).^4^ Furthermore, the gender composition of siblings can put later-born daughters at a disadvantage through two mechanisms–sibling rivalry and the fertility-stopping effect (Jayachandran and Pande, 2017).^5^ We specifically examine the impact of the HSAA on daughters’ health and later-born children. We find no evidence that the HSAA led to improvements in the health of daughters and later-born children.

We also examine the several channels that are consistent with the positive health effect of the HSA in data. Specifically, we examine the effect of the HSAA policy on several proxies of female empowerment: women’s say about household large purchases, women’s say about her health care choices, and women can travel to market alone. We show evidence that the HSAA amendments led to a robust increase in female bargaining. The theoretical model underlying predictions based on a non-unitary household (Browning et al., 1994) shows that an increase in female bargaining power is likely to improve child height. This argument is consistent with extensive literature that has tested whether income in the hands of women of a household has a different impact on intra-household allocation than income in the hands of the men (Duflo, 2012).

Our study contributes to the existing empirical literature on how female empowerment can improve economic outcomes in low-and middle-income countries (LMICs). First, we shed light on how an inheritance-related policy, the HSAA^6^, can affect child height in the context of a developing country. The causes of child stunting are numerous, ranging from genetics, poor nutrition to repeated disease insults. An inadequate diet lacking in calories, protein, or other micronutrients at early ages can affect children’s growth. We show clear evidence that female empowerment can exert a significant influence on child height. Previous research shows that children’s outcomes improve when women have more control in the household (Duflo, 2003; Lundberg, Pollak and Wales, 1997; Qian, 2008). We add to this literature by showing that one key channel via which female empowerment can improve children’s outcomes is via improved inheritance rights standing. Second, although female empowerment can improve children’s height in the context of a developing country, we also detect cautionary evidence that improvements in female bargaining power do not necessarily translate to better human capital outcomes for all children. Our findings imply that important gender and birth order considerations likely play an essential role as we find no evidence that the HSAA led to improvements in the health of daughters and later-born children.

The rest of the paper is structured as follows. Section II provides background on the Hindu Succession Act and evidence on the Act’s impact on female bargaining. Section III describes the data. Section IV presents the empirical strategy. Section V presents the results, and Section VI concludes.

## II. Background

### A. The Hindu Succession Act 1956, the 2005 Amendment, and the Impact on Women’s Bargaining

In a series of sweeping legal reforms in the 1950s, India introduced the Hindu Succession Act (*HSA*) in 1956. The main objective of the legislation was to unite two existing inheritance systems in the country. The Act also aimed to clarify the inheritance rights of women over private properties.

Before 1956, two different systems guided property inheritance in India: *Dayabhaga* (in West Bengal and Assam) and *Mitakshara* (the rest of India) (Agarwal 1994). From the early twelfth century and until the Hindu Succession Act (HSA) enactment in 1956, two principal systems guided inheritance procedures within Hindu communities in India. One was the Dayabhaga, and the other was the Mitaksara. The Dayabhaga, written by the Indian Sanskrit Scholar Jimutavahana, was a Hindu law treatise. The treatise primarily focused on rules regarding inheritance procedures within the Hindu community. The Dayabhaga was the most decisive authority in Modern British Indian courts in the Bengal region of India. The second tradition, the Mitaksara, guided the legal system in the rest of the country. The Mitakshara (which translates to *measured words*) is regarded as an authority even in Bengal in all legal matters if no conflicting opinions exist within the Dayabhaga.

The main distinction between these two systems was how they categorized and distinguished between private (separate) and any joint family (or ancestral^7^) property. Private property is generally self-acquired and cannot pass via patrilineal succession (i.e., from a male line, such as a father, a grandfather). Ancestral or joint family property, on the other hand, is commonly inherited patrilineally. The *Dayabhaga* system did not distinguish between the two types of properties. Under the *Dayabhaga system,* all heirs, including sons, daughters, or widows, could claim over inheriting property. By comparison, the *Mitakshara* system distinguished the inheritance procession depending on whether the property was considered private or joint-family property. Under this system, the owner can bequeath any privately owned to anyone he wished. However, only *coparceners* (i.e., sons, grandsons, or great-grandsons) could inherit joint family property.

The HSA aimed to promote gender equity by conferring equal rights over private property to women. However, the Act did not apply to the joint family property. Therefore, under the HSA, daughters had an equal right to their father’s private property if the Hindu (i.e., Hindus, Buddhists, Jains, and Sikhs) male died without a will^8^ *(intestate)*. However, women could not inherit joint family property. In contrast, sons being coparceners had the right to inherit the joint family property by birth which implied that their share of the property could not be willed away. They could even demand a division of the joint family property in case older coparceners are alive. Therefore, and because of its different treatment of men and women who could inherit joint family property, the HSA discriminated against women.

Both the federal and the state governments had legislative authority over inheritance issues in India (Roy, 2015).^9^ Although HSA was a federally mandated law, states could pass amendments to it, and these amendments guided the rules within the jurisdictions of these states. For instance, Kerala^10^ (in 1976), Andhra Pradesh (in 1986), Tamil Nadu (in 1989), Karnataka (in 1994), and Maharashtra (in 1994) passed state amendments to the HSA. These amendments provided Hindu women equal inheritance rights over the joint family property so long as they were not married at the time of the amendment^11^.

The HSAA, in the *Mitakshara* system, effectively elevated the status of daughters to that of sons, but these benefits pertained to unmarried daughters. Daughters who married before the commencement of the HSAA cannot claim coparcenary status in the HSAA. This aspect of the law implies that married daughters do not enjoy the same rights regarding a coparcenary property as the sons. There are three reasons why the law excluded married daughters. First is the dowry practice. Second, in some communities (such as *Kammas*), daughters receive a share of the property at the time of marriage. However, the dowry practice is technically illegal in India. Moreover, only a few communities practice giving property at the time of marriage. Therefore, a blanket exclusion of married daughters, based on an illegal act and practice in a few exceptional communities, cannot be a reasonable justification (Sivaramayya, 1988). Finally, there is a cultural mindset that the daughter becomes part of the husband’s family, and this mindset plays a significant role.

In 2005, India passed a federal amendment, extending the HSA to the entire country regarding the gender disparity over inheritance rights. In particular, the amendment targeted and modified Section 6, which had initially provided the basis for gender discrimination on who could claim inheritance rights over joint family property. After the passing of the amendment in 2008, the Supreme Court ruled that the law had a retrospective effect. Under this effect, a daughter could become a co-sharer with her male siblings, and the father would not have had to be alive as of 9 September 2005 (when India passed the amendment).

### B. Child Health in India

Child height is a significant predictor of adult human capital and better economic outcomes (Case and Paxson, 2010). Existing research has also documented the role of several crucial channels, such as better adult physical health (Case and Paxson, 2010), cognition (Case and Paxson, 2008), social dominance (Hensley, 1993), discrimination (Hamermesh and Biddle, 1994), and the social repercussions of being short in adolescence (Persico et al., 2004).

Despite India’s economic progress in the last few decades, health outcomes are considerably worse than predicted based on international comparisons with other developing countries (Drèze and Sen, 2013). Because of the robust empirical evidence regarding the critical lifetime influence child health can exert, child height in India has received particular attention in the economics literature. The average child under 5 in rural India is about two standard deviations shorter than the World Health Organization (WHO) reference population for healthy growth. In 2020, 31 percent of Indian children under five remained stunted (WHO 2021). Thus, despite a GDP per capita higher than that of over 60 countries, India has the fifth-highest stunting rate in the world (Krishna et al., 2017). The average child born in India is more likely to be stunted than her counterpart in Sub-Saharan Africa, even though their mother has higher birth survival; on average, the parents are more affluent and more educated (Gwatkin et al., 2007).

There is also growing empirical evidence regarding the crucial role parental investments play in determining child height in India (Jayachandran and Pande, 2017). A critical factor for lower height entails lower parental investments relates to differential parental investment based on gender and birth order. For instance, Borooah (2004) shows evidence of pro-boy parental investments (in dietary diversity) in children aged up to 24 months born. DasGupta (1987) shows that although infant girls and boys obtain the same caloric intake, families feed girls more cereals while giving boys more milk and fats. Fledderjohann et al. (2014) also report higher chances of milk consumption for under-five boys than females.

However, enhancing female empowerment at the household level and policy initiatives to improve female bargaining power can positively influence child health. When women are decision-makers within the household, how much they bring to the table can impact ultimate choices. Empirically, an extensive literature has tested whether income in the hands of women has a different impact on intra-household allocation than income in the hands of the men (Duflo, 2012). The evidence suggests that, compared to income or assets in the hands of men, income or assets in the hands of women is associated with more significant improvements in child health and larger expenditure shares of household nutrients (Thomas, 1990, 1992). The theoretical model underlying these predictions is of a non-unitary household, a household as a collective of individuals with different preferences (Browning et al., 1994).

In India, a predominantly rural country, one crucial way to enhance female empowerment relates to inheritance rights for the land and family property. Land ownership is a critical determinant of economic and social status (Agarwal, 1994), and inheritance is the primary way to acquire it. However, conventional male-biased inheritance practices and cultural conservatism (Agarwal, 1994) engender limited earning opportunities and bargaining power for married women in India. Because of the potential to transform women’s inheritance rights, some Indian states amended the male-favored Hindu Succession Act (HSA) by providing married women an equal share in ancestral property. As a result, improved female bargaining power could improve human capital outcomes and gender inequality.

In developing countries such as India, high levels of human capital may offer a way to escape poverty (Chakravarty et al., 2018). Therefore, policies aimed to improve female empowerment can create positive spillover effects for children. Although there is evidence that some public policies can be highly effective, their design and targeting are crucial for human capital accumulation. For example, the provision of improved inheritance rights for women can improve children’s human capital outcomes. It is this empirical possibility that we attempt to shed light on our analyses.

## III. Data and Summary Statistics

### A. The Data: A Descriptive Analysis

For our analysis, we use data from the Indian National Family and Health Survey (NFHS), a cross-sectional survey comprising three rounds of data: Wave 1 (1992-1993), Wave 2 (1998-1999), and Wave 3 (2005-2006). The household survey covers rural and urban households. It adopts a stratified multistage cluster sampling method to identify a nationally representative sample of the population living in both urban and rural areas in 29 states. The survey selected 110,000 households in each wave and collected information from 125,000 women (aged 15 to 49 years) and 75,000 men (aged 15 to 54 years).

All women in the age range were eligible for an interview. However, because numerous health indicators pertained to the sample of ever-married women and children, the required sample size for men was considerably smaller. Thus, of the 216,969 eligible women and men, 124,385 women and 74,369 men participated in the survey, yielding a response rate of 94.5 percent and 87.1 percent, respectively.

The survey questionnaire comprises several distinct modules, including a household module, a module collecting information from women, and a village information module. The household module collected information from face-to-face interviews. It draws information from all residents in each sample household. In addition, the survey covers demographic information on age, gender, marital status, relationship to the head of the household, education, and occupation for each listed person. Based on the household module, the survey team identified respondents eligible for the woman’s questionnaire, which collected additional demographic information on adult female respondents and their children (if any).

The woman’s questionnaire collects information from all ever-married women aged 15–49 who were the residents of the sampled household. The module collects background information on socioeconomic characteristics (age, marital status, education, employment status, place of residence), reproductive behavior (fertility choice, birth spacing, number of children, prenatal and postnatal healthcare use), and quality of care. The module also covers questions on all children (their age, sex, birth order, and health information, such as height, weight, hemoglobin level, and prior vaccinations). The survey gathered anthropometric measures for both adults and children. The NFHS collected data on height and weight measurements for children in all rounds. Height data for adults is available only in NFHS-2, NFHS-3, and NFHS-4.

Our analysis sample consists of 67,815 children for whom we have anthropomorphic data. We use data on the following fifteen states: the four treatment states are Andhra Pradesh, Tamil Nadu, Maharashtra, and Karnataka; and the control states are Arunachal Pradesh, Bihar, Goa, Gujrat, Haryana, Himachal Pradesh, Madhya Pradesh, Orissa, Panjab, Rajasthan, and Uttar Pradesh.^12^

### B. Construction of Study Outcomes

#### Child Health

We use height as an indicator of early childhood health. Height is a stock variable and meaningful indicator of accumulated decisions regarding nutritional intake in early life (Case and Paxson, 2010). We also use weight as an additional proxy of child health, although weight is a flow measure and captures short-term changes to the nutritional environment.

Based on the raw measurements for these variables, we create standardized measures for height and weight using additional information on age, height, and weight for all children. The two standardized measures for height and weight are height-for-age (HFA) and weight-for-age (WFA).^13^ The HFA z-score is available for children under age five. The WHO defines children who are two and three standard deviations lower than the mean HFA z-score as moderately and severely stunted (WHO, 2006). Similarly, children who are two and three standard deviations lower than the mean WFA z-score are defined as moderately and severely wasted, respectively.

We also examine other health indicators of child health. We use indicators, such as total prenatal visits, whether mother took iron supplementation, tetanus shot, whether the delivery was done at a health facility, whether there was a postnatal check within two months of birth, and whether the child was ever vaccinated. Postnatal checks are only available only for the youngest living child. The recommended vaccination age is up to 12 months, so we restrict the sample for total vaccinations to children ages 13 to 59 months. We also create a composite normalized input index based on the prenatal and postnatal inputs. This index helps us gauge the parental investment care for children. Furthermore, we measure disease incidence in the last two weeks to capture the early childhood disease environment. We do so by constructing a composite index based on the following variables: the incidence of the child having a fever in the last two weeks, the incidence of cough in the last two weeks, and the incidence of diarrhea in the last two weeks. Based on these variables, we create a normalized composite score.

#### Female Bargaining Power Measures

In addition to the socioeconomic and health outcomes in the survey, we also explore outcomes related to childbearing and women’s autonomy. The IDHS collects data on the number of children ever born and on the mother’s age at first marriage. We measure women’s bargaining by using three binary variables: whether the woman has a say about large purchases, whether the woman has a say about her own healthcare choices, and whether she can go to market alone. This approach followed the methodology adopted by Heath and Tan (2020). Based on the individual indicators, we generate a composite normalized index for female bargaining power.

### C. Sample Summary Statistics

Table 1 reports the summary statistics for the main variables. The table reports data on the entire sample, the sample in states affected by the policy change (Columns 2 through 4), and the sample only in states unaffected by the policy (Columns 5 through 7). The policy change primarily affects the patrilineal and patrilocal kinship who are Hindus. Therefore, we split the sample by religion: column 2 comprises the Hindus affected by the policy; column 3 reports data only on non-Hindu individuals in the treated states. Columns 5-7 presents the summary statistics for the sample (Hindus and non-Hindus) and non-treated states. Table 1 shows data based on the mothers’ sample: age, age at first marriage, age at first birth, and various proxies of bargaining power.

**Table 1:** Summary Statistics

|  | Full Sample | Treated States |  |  | Non-Treated States |  |  |
| --- | --- | --- | --- | --- | --- | --- | --- |
|  |  | Treated religion | Non treated religion | All | Treated religion | Non treated religion | All |
|  | (1) | (2) | (3) | (4) | (5) | (6) | (7) |
| Household have electricity (1=if yes) | 0.633<br>(0.482) | 0.823<br>(.382) | 0.890<br>(0.313) | 0.836<br>(0.370) | 0.578<br>(0.494) | 0.779<br>(0.415) | 0.579<br>(0.494) |
| Household receive piped water at home (=1 if yes) | 0.315<br>(0.465) | 0.504<br>(0.50) | 0.444<br>(0.497) | 0.530<br>(0.499) | 0.247<br>(0.431) | 0.470<br>(0.499) | 0.258<br>(0.437) |
| Mother's age (years) | 26.11<br>(5.40) | 24.81<br>(4.72) | 26.52<br>(5.33) | 24.96<br>(4.84) | 26.22<br>(5.38) | 28.97<br>(5.59) | 26.41<br>(5.50) |
| Mother's age at first marriage (years) | 17.36<br>(3.28) | 17.60<br>(3.51) | 20.34<br>(4.50) | 17.65<br>(3.52) | 17.24<br>(3.16) | 20.96<br>(4.71) | 17.28<br>(3.21) |
| Total children born to a mother | 3.027<br>(1.911) | 2.383<br>(1.372) | 2.00<br>(1.101) | 2.461<br>(1.458) | 3.081<br>(1.904) | 2.781<br>(1.917) | 3.179<br>(1.988) |
| Say about large purchase (=1 if yes) | 0.496<br>(0.5) | 0.515<br>(0.5) | 0.569<br>(0.496) | 0.513<br>(0.5) | 0.48<br>(0.5) | 0.704<br>(0.457) | 0.491<br>(0.5) |
| Say about own health (=1 if yes) | 0.549<br>(0.498) | 0.569<br>(0.495) | 0.636<br>(0.482) | 0.57<br>(0.495) | 0.531<br>(0.499) | 0.68<br>(0.467) | 0.543<br>(0.498) |
| Go to market alone (=1 if yes) | 0.409<br>(0.492) | 0.548<br>(0.498) | 0.602<br>(0.49) | 0.533<br>(0.499) | 0.361<br>(0.48) | 0.694<br>(0.461) | 0.371<br>(0.483) |
| Bargaining | 0.00<br>(1.00) | 0.137<br>(0.99) | 0.295<br>(0.987) | 0.125<br>(0.994) | -0.067<br>(0.996) | 0.542<br>(0.894) | -0.038<br>(0.999) |
| Child is a girl (=1 if yes) | 0.481<br>(0.5) | 0.476<br>(0.499) | 0.506<br>(0.501) | 0.479<br>(0.5) | 0.479<br>(0.5) | 0.494<br>(0.5) | 0.481<br>(0.5) |
| Child's birth order | 2.798<br>(1.896) | 2.182<br>(1.362) | 1.836<br>(1.067) | 2.250<br>(1.446) | 2.855<br>(1.892) | 2.568<br>(1.871) | 2.945<br>(1.974) |
| Child's HFA z-score | -1.918<br>(1.851) | -1.665<br>(1.737) | -1.231<br>(1.679) | -1.659<br>(1.738) | -1.985<br>(1.86) | -1.236<br>(1.874) | -1.988<br>(1.874) |
| Child's WFA z-score | -1.788<br>(1.377) | -1.663<br>(1.293) | -1.28<br>(1.236) | -1.639<br>(1.297) | -1.835<br>(1.391) | -1.179<br>(1.411) | -1.828<br>(1.395) |
| Childhood Disease Environment Index | 0.00<br>(1.00) | -0.060<br>(0.946) | 0.007<br>(0.985) | -0.052<br>(0.957) | 0.004<br>(1.003) | 0.048<br>(0.992) | 0.014<br>(1.011) |
| Number of children | 67,815 | 9,198 | 394 | 11,163 | 31,211 | 702 | 36,691 |

At the time of first marriage, in both treated and non-treated states, the average mother’s age is around 17. However, non-Hindu mothers in all states, regardless of whether they reside in areas affected by the policy, marry at a slightly older age (i.e., 20 years).

However, Hindu mothers living in areas affected by the policy have a lower-than-average age at first marriage and a higher-than-average age when they have their first. Overall, and based on the index we construct, Hindu women have lower bargaining power than non-Hindu women. However, Hindu women in states affected by the HSA exhibit higher bargaining power than Hindu women in the states that did not adopt the HSA.

Turning to the children’s characteristics in the whole sample, Table 1 reports the average age to be around two years. As a proportion of all children, the share of daughters is nearly identical between states who adopted the HSA or not. The average HFA is -1.918, but it is lower in states that adopted the HSA (i.e., -1.659); in states that did not, the HFA is - 1.988. The summary statistics for the WFA measure present the same pattern. Finally, Hindu children in non-treated states show a clear disadvantage in prenatal and postnatal health inputs. They receive lower parental inputs than Hindu children in treated states.

## IV. Empirical Strategy

To estimate the effect of the HSA amendment on child health outcomes, we take advantage of the staggered adoption of the HSA across states. We use two sources of identifying variation. The first source is the timing of a woman’s marriage. The second source relates to whether the woman’s state of residence adopted the 1956 HSA; the reform only affects unmarried women when the reform occurred in their state^14^; women who were married at the time of the reform form the control group. Four states – Andhra Pradesh, Tamil Nadu, Karnataka, and Maharashtra – adopted the Act. Eleven states did not: Arunachal Pradesh, Bihar, Goa, Gujrat, Haryana, Himachal Pradesh, Madhya Pradesh, Orissa, Panjab, Rajasthan, and Uttar Pradesh.

To study the impact of the HSA on child health, we employ a difference-in-differences strategy similar to the approach used by Duflo (2001) and by Lavy and Zablotsky (2015), i.e., we rely on differences across cohorts and groups affected or not by the policy reform. We estimate the effect of the HSA amendments using the following reduced-form specification:

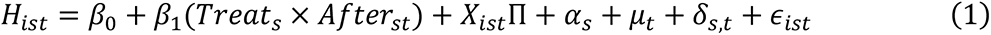

*H*_*ist*_ is the outcome for child *i* born to a mother, who married in year *t*, in state *s*. *Treat* is a dummy variable, and it equals one if the mother is from a state that amended HSA before the national adoption in 2005 (i.e., reform states) and equals zero if not from one of the reform states. *After*_*st*_ is a dummy variable: it takes a value of 1 if the mother was married after the reform in her state *s*, and takes zero otherwise. *Treat* × *After*_*st*_ is a binary indicator, set to 1 if the mother is from a reform state *s* and was married after the reform in her state *s*, and equals 0 if she is not exposed to the reform. ***a***_*s*_ is state fixed effect, *μ*_*t*_ is the mother’s year of birth fixed effect, and *δ*_*s*,*t*_ captures state-year fixed effects. State fixed effects capture state-specific characteristics; the year fixed effect accounts for time-varying but group-invariant factors. Finally, the state-year fixed effect allows us to control state-specific time-varying omitted variables, which may correlate with the HSA amendment. ***X***_***i***_ is a set of mother-related characteristics (age at first birth, age at first birth squared, age); it also includes child characteristics, such as gender, age, and birth order.

The parameter of interest, *β*_1_, captures the effect of the HSA. The validity of our empirical approach depends on two assumptions: (a) no pre-trends exist for the treatment and control groups, and (b) states do not adopt the HSA amendment in a manner correlated with child health.

Before we proceed with estimating (1), we examine the trend in the outcome variables for the treated and non-treated areas. Figure 1 displays the trajectory of the HFA and WFA outcomes. For both outcomes, and regardless of the treatment status, the figure reveals an upward trend. However, HFA in the treated states is higher than in the non-treated states between 1978 and 2004. On the other hand, the WFA is quite similar between the treated and non-treated states until 1999; the gap widens after that.

**Figure 1:**
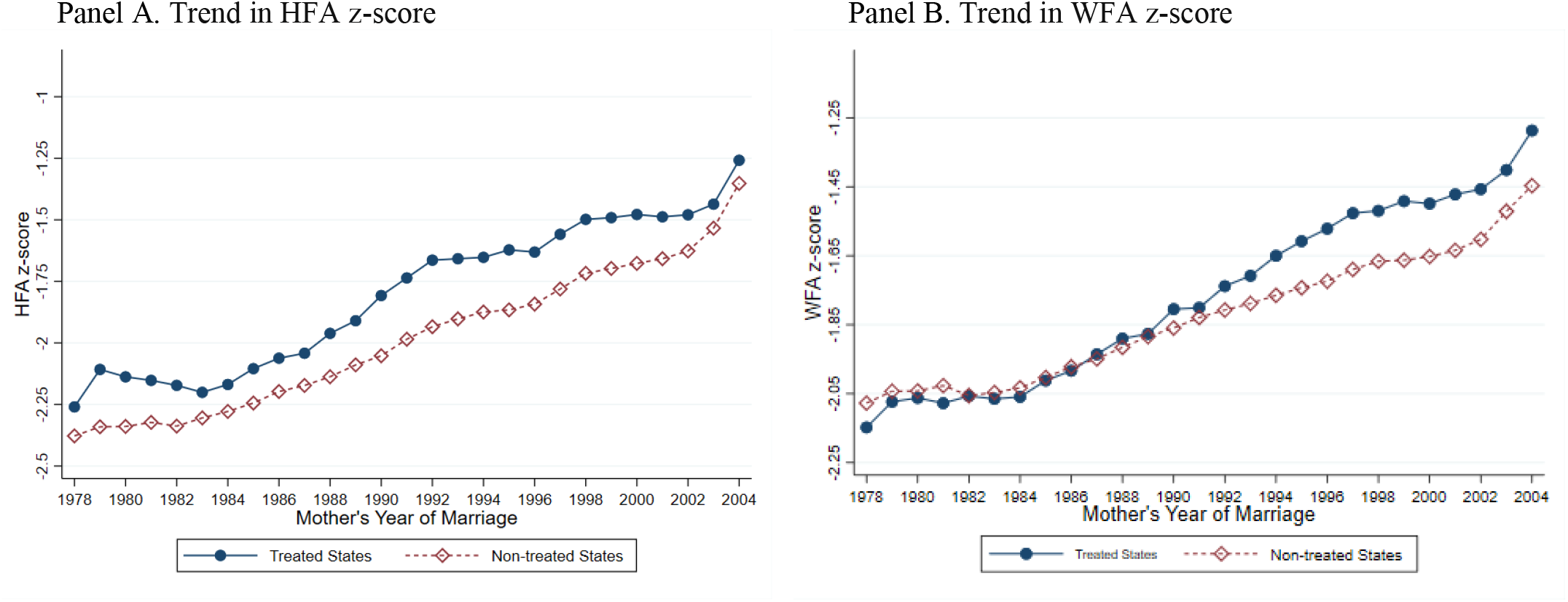
Trends in Child Health Outcomes in Treated and Non-Treated Areas.

Except for Maharashtra, the HSAA reforms are in southern Indian states. These states tend to outperform the rest of the country in terms of child health. As noted earlier in Table 1, the treated areas exhibit better child health indicators than the non-reform states. This discrepancy could be a cause of concern for our empirical strategy.

We conduct an exercise to examine the similarity between the trends of the health indicators. Figure 2 displays the trajectory of the average HFA and WFA measures. The figure visually compares the evolution of health measures in the reform and non-reform states before enacting the HSAA policies. Because the reforms happened in different years, we include all reform states up to 1984. For example, in 1985, Andhra Pradesh’s reform was enacted. Therefore, we drop it from the comparison group. Likewise, Tamil Nadu enacted the reform as a law in 1989. Therefore, we drop the state for 1989 only in this particular data exercise. Although the average of the health indicators is higher in the reform states, the trend in the HFA and WFA measures is similar. Thus, the overall pattern does not provide visual justification for differential trends between the two groups.

**Figure 2:**
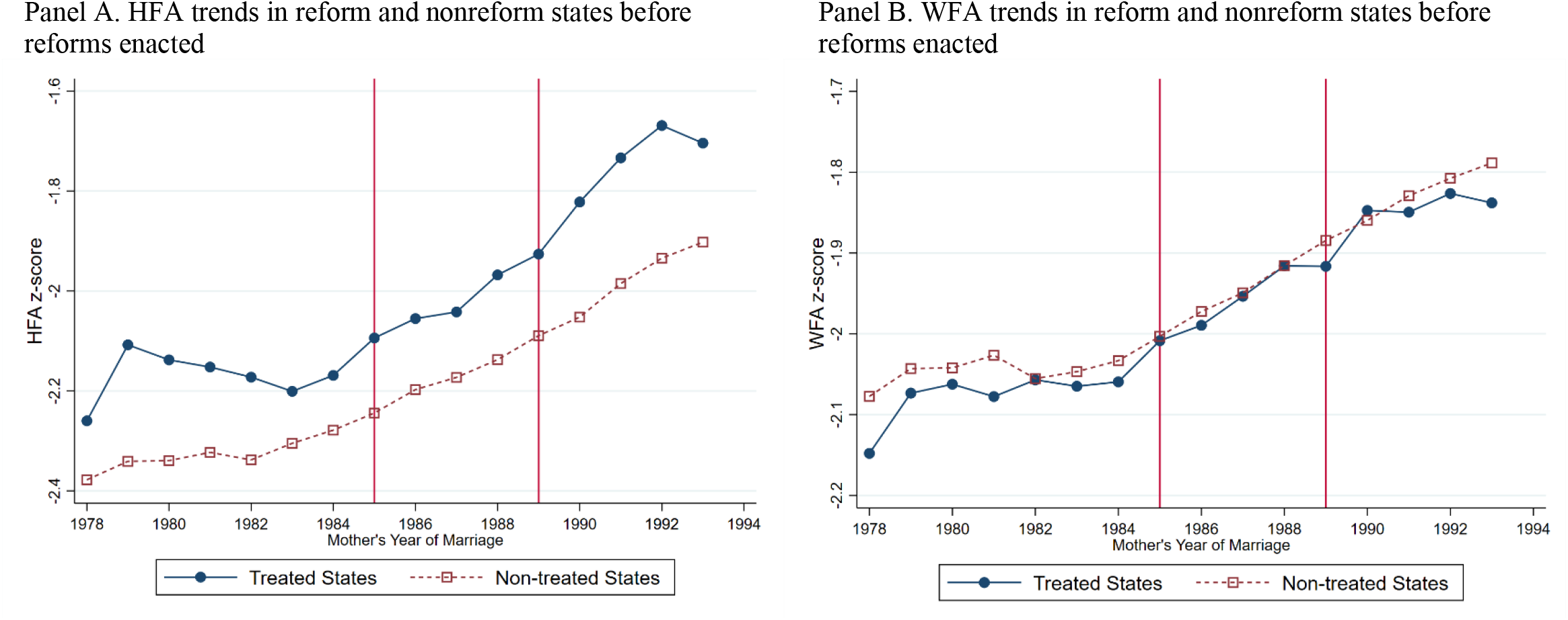
HFA and WFA Trends in Reform and Non-reform States Before the Enacted Reforms

Next, we formally test whether there is a similar evolution pattern of child health in the treated and non-treated states before the reform using a regression approach described in Angrist and Pischke (2009). In this exercise, we use data from both treatment and control states before the actual reform. Specifically, we use data until 1984 as the first state amended the HSA in 1985. We estimate:

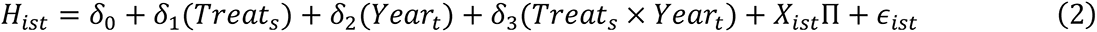

where *H*_*ist*_ is the outcome for child *i* born to a mother, who married in year *t*, in state *s*. *Treat* is a dummy variable, and it equals one if the mother is from a state that amended HSA before the national adoption in 2005 (i.e., reform states) and equals zero if not from one of the reform states. *Year*_*t*_ is a linear trend of mother’s year of marriage runs from 1970 to 1984. ***X***_***i***_ is the same set of controls as in equation (1). *β*_3_ captures the differential trend in the outcome variable between treated and non-treated states before the HSA amendment.

We report the results for this exercise in Table 2. The coefficients of Treat × Year in columns 1 and 3 are insignificant and close to zero for both HFA and WFA. The inclusion of controls (reported in columns 2 and 4) only strengthens these results. Therefore, this exercise further bolsters the validity of our empirical approach.

**Table 2:** Test of Common Trend

|  | HFA z-score |  | WFA z-score |  |
| --- | --- | --- | --- | --- |
|  | (1) | (2) | (3) | (4) |
| Treat x Year | 0.011<br>(0.014) | 0.008<br>(0.014) | 0.007<br>(0.011) | 0.008<br>(0.011) |
| Treat | -0.051<br>(0.179) | 0.126<br>(0.171) | -0.187<br>(0.135) | -0.131<br>(0.137) |
| Year | 0.017***<br>(0.006) | 0.056***<br>(0.009) | 0.007*<br>(0.007) | 0.043***<br>(0.008) |
| Observations | 10,331 | 10,330 | 10,226 | 10,224 |
| Survey year FE | Yes | Yes | Yes | Yes |
| Controls | No | Yes | No | Yes |

Note: (a) Treat is a dummy variable equals 1 if the state amended the HSA and equals zero otherwise. (b) Year is a linear trend of mother's year of marriage (c) HFA z-score is the Child's height-for-age z-score, and WFA z-score is child's weight-for-age z score. (d) Standard errors are clustered at the PSU level and appear in parentheses. (e) Survey year FE are added in all columns. (f) Control variables are: mother's age at first birth, mother's age at first birth squared, mother's current age dummies (15-20 years, 21-25 years, 26-30 years, 31-35 years, and 35+ years), child's sex, child age dummies (months), and child's birth order (2nd born, and 3rd+ born). (g) The sample used in this table is Hindus who are subject to HSA 1956.

## V. Results

### A. Effect of the Reform on Child Health

Table 3 reports our main results, which are the estimates based on equation (1). The dependent variables are the primary health outcomes and the parental investment in child health. Our variable of interest, *Treat* × *After*, is a dummy capturing whether the mother is from a reform state *s* and was married after the reform in her state *s*. The table contains the OLS coefficients and standard errors in parentheses. We cluster the standard errors at the primary sampling unit (PSU). All columns of Table 3 include control variable and fixed effects (FE)–state fixed effect, year (mother’s year of birth) fixed effect, state-year fixed effect, and survey year fixed effect.

**Table 3:** Effect of HSAA on Child Health

|  | HFA z-score | WFA z-score | Prenatal Inputs |  |  | Postnatal Inputs |  |  | Pooled inputs |
| --- | --- | --- | --- | --- | --- | --- | --- | --- | --- |
|  |  |  | Total prenatal visits during pregnancy | Mother took iron supplement during pregnancy (1= if yes) | Mother took tetanus shot during pregnancy (=1 if yes) | Delivery at health facility (1= if yes) | Postnatal check within two months of birth (1= if yes) | Ever vaccinated (1= if yes) |  |
|  | (1) | (2) | (3) | (4) | (5) | (6) | (7) | (8) | (9) |
| Treat × After | 0.183***<br>(0.065) | 0.276***<br>(0.049) | 0.805***<br>(0.119) | -0.013<br>(0.016) | 0.026*<br>(0.014) | 0.173***<br>(0.018) | 0.139***<br>(0.026) | -0.064***<br>(0.010) | 0.359***<br>(0.048) |
| Observations | 56,832 | 56,477 | 50,338 | 33,444 | 33,482 | 56,796 | 23,758 | 56,797 | 23,633 |
| State FE | Yes | Yes | Yes | Yes | Yes | Yes | Yes | Yes | Yes |
| Year FE | Yes | Yes | Yes | Yes | Yes | Yes | Yes | Yes | Yes |
| State-Year FE | Yes | Yes | Yes | Yes | Yes | Yes | Yes | Yes | Yes |
| Survey year FE | Yes | Yes | Yes | Yes | Yes | Yes | Yes | Yes | Yes |
| Controls | Yes | Yes | Yes | Yes | Yes | Yes | Yes | Yes | Yes |

The estimates of the child health outcomes–height-for-age (HFA) z-score and weight-for-age (WFA) z-score–are presented in columns 1 and 2. The coefficients of *Treat* × *After* in columns 1 and 2 are positive and statistically significant. For example, in column 1, the coefficient of *Treat* × *After* is 0.183, which indicates that the HSA reform has on average increased children’s HFA z-score by 0.183 standard deviations. Similarly, the coefficient of *Treat* × *After* in column 2 indicates that the HSA reform has, on average increased children’s WFA z-score by 0.276 standard deviations. These results suggest that women favored inheritance rights reform in India has significantly improved child height and weight.

Table 3 also shows whether the HSA amendment leads to increased parental investment in children’s health. Outcome variables in columns 3-6 are prenatal inputs, and outcome variables in columns 7-8 are postnatal inputs. We have four indicators of prenatal inputs during pregnancy–total prenatal visits, mother took iron supplementation, the mother took tetanus shot, and delivery at a health facility. Delivery at a health facility indicates whether the childbirth happened at a health facility instead of birth at home. Total prenatal visits and delivery at a health facility are available for all children under five years.

In contrast, data on iron supplement intake and tetanus shot is available only for the most recent birth (youngest child). We have two indicators of postnatal parental inputs–postnatal check within two months of labor and child ever vaccinated. Postnatal check within two months of birth is available only for the most recent delivery (youngest child). Finally, the outcome variable in the last column is pooled inputs, which the first principal component of the standardized prenatal and postnatal inputs, and then normalized to have mean zero and standard deviation one.

Columns 3 and 9 show the OLS coefficients of *Treat* × *After*, and columns 4-8 show the coefficients of the linear probability model. The coefficients of *Treat* × *After* in columns 3-6 are positive and statistically significant, except iron supplement intake in column 4. The coefficient in column 3 shows that the HSA amendment leads to about 0.81 higher prenatal visits during pregnancy. Similarly, the HSA amendment has increased the probability of a mother taking a tetanus shot by 2.6 percent and delivering at a health facility by 17.3 percent. The coefficients of *Treat* × *After* for postnatal visits within two months is 0.139, suggesting that the HSA amendment has increased the likelihood of postnatal check within two months of birth by about 14 percent. Although the coefficient associated with the person being ever vaccinated is negative, the overall pooled input corroborates the significant positive effect of the HSA amendment. The coefficient of *Treat* × *After* in column 9 indicates that the amendment has increased overall parental investment in children by about 36 percent.

We examine whether there is an upward trajectory of the health outcomes (HFA or WFA) in correspondence with the area adopting the HSAA. For this exercise, we estimate equation (1) substituting the HSAA policy variable with a complete set of dummy variables going from 10 years before adopting the HSAA to 10 years after. We display the results in Figure 3. The figure displays the coefficients associated with the dummy variable in a specification where the outcome variables are the two primary outcomes, the HFA and the WFA measures. Figure 3 shows no evidence that the improvement in the health outcomes occurred before the HSAA enactment. None of the displayed coefficients for the years prior or the years following the HSAA enactment in a particular state are significantly different from zero. This exercise bolsters the results reported in Table 3, as it will be challenging to account for the improvements of the health outcomes in the years immediately following the HSAA enactments.

**Figure 3:**
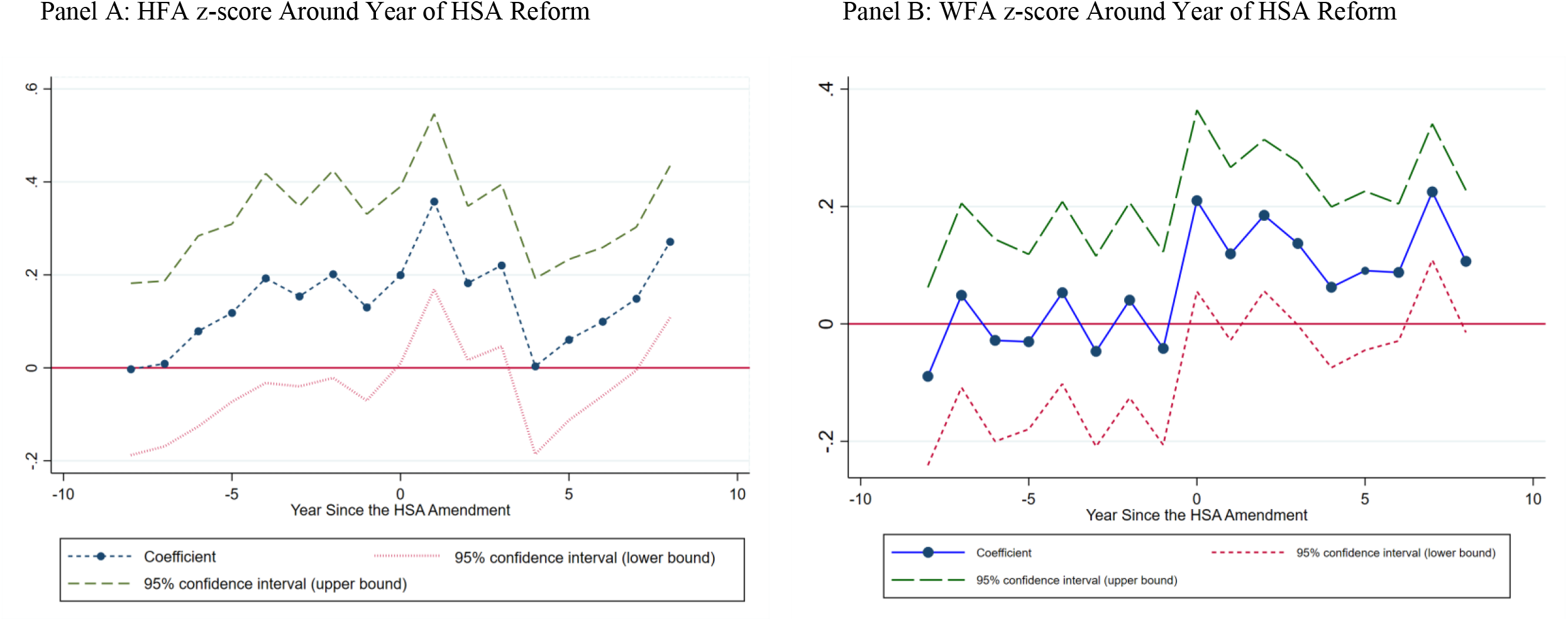
Health Indicators Around the Year of HSA Reform

Another way to investigate the possible selection into the HSAA is to formally test for any correlation between the HSAA and average child health before the amendments. The earliest survey years available for child health is from the NFHS-1 for the 1992-1993 period. Given that our objective is to examine the relationship between the HSAA and health outcomes at the baseline period, we focus on the states that adopted the HSAA after 1993: Maharashtra (1994) and Karnataka (1994). Unlike the subsequent survey waves used in the primary analysis, data in this particular wave is also available at the district level. We analyze the health outcomes for Hindu-only households at the district level. We estimate the following equation.

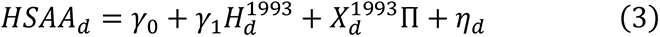

where *HSAA*_*d*_ is a dummy variable equals one if the district is subject to HSA amendment after 1993 and before the national adoption in 2005, and equals zero otherwise; 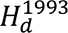 is the child health outcomes–HFA and WFA z-scores–in district *d* in the year 1992-93; 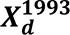 is the vector of controls capturing the average of various socioeconomic characteristics for the district in 1992-93 (i.e., mother’s age at first birth, mother’s age at first birth squared, mother’s current age dummies). We estimate equation (3) twice for the HFA and WFA outcomes. Table 4 reports the results.

We first regress the district’s HSAA status on the average HFA z-score in 1992-93, without any other controls (column 1 of Table 4 reports the results). The coefficient of HFA is close to zero and statistically insignificant. The inclusion of controls in column 2 does not alter the result: we find no evidence of a correlation between the HSAA adoption and HFA outcomes. We obtain similar results for WFA outcome (at the district level), thereby bolstering our claim regarding the absence of a relationship between the HSAA and both child health outcomes.

**Table 4:** Possible Selection in HSA Amendment

|  | Outcome variable: HSA Amendment (=1 if yes) |  |  |  |
| --- | --- | --- | --- | --- |
|  | (1) | (2) | (3) | (4) |
| Average levels in Districts in 1992-93 |  |  |  |  |
| HFA | 0.026<br>(0.113) | 0.024<br>(0.059) | - | - |
| WFA | - | - | -0.077<br>(0.115) | -0.049<br>(0.070) |
| Observations | 217 | 217 | 217 | 217 |
| Controls | No | Yes | No | Yes |

*Note:* (a) Sample used in this table are Hindus (who are subject to HSA 1956) from the NFHS-I (1992-93) survey and states that do not amend HSA until 1993. (b) HSA reform is a dummy variable that equals one if a district is subject to the HSA amendment after 1993 (i.e., Karnataka (in 1994) and Maharashtra (in 1994) and equals zero otherwise. (c) HFA refers to the mean childrens' height-for-age z-score at the district level, and WFA refers to the mean children's weight-for-age z-score at the district level. (d) Bootstrapped standard errors appear in parentheses. Standard errors are bootstrapped to address the small sample issue. (e) Control variables are: mother's age at first birth, mother's age at first birth squared, mother's current age dummies (15-20 years, 21-25 years, 26-30 years, 31-35 years, and 35+ years). (f) \*\*\*Significant at the 1 percent level, \*\*Significant at the 5 percent level, \*Significant at the 10 percent level.

### B. Placebo Exercises

We conduct a falsification exercise to examine the validity of our estimation approach. Specifically, we re-estimate our estimation equation (1) model but among the sample of Christians. Since the HSA amendment affects only the Hindu population, this particular placebo exercise should produce no effects of the HSA for the Christian sample. Table 5 reports the results based on this placebo test. The coefficient of interest, based on the OLS estimation, is the estimate for the *Treat* × *After* variable. The associated estimates for HFA and WFA outcomes are close to zero and statistically insignificant. Similarly, the linear probability model coefficients (reported in columns 4-8) and the OLS coefficient (reported in column 9) are close to zero and statistically insignificant. Therefore, the placebo exercise bolsters our primary estimation method and is evidence that our estimation technique does not pick up spurious effects on child health outcomes.

**Table 5:** Placebo Treatment Effect of HSAA on Child Health

|  | HFA z-score | WFA z-score | Prenatal Inputs |  |  | Postnatal Inputs |  |  | Pooled inputs |
| --- | --- | --- | --- | --- | --- | --- | --- | --- | --- |
|  |  |  | Total prenatal visits during pregnancy | Mother took iron supplement during pregnancy (1= if yes) | Mother took tetanus shot during pregnancy (=1 if yes) | Delivery at health facility (1= if yes) | Postnatal check within two months of birth (1= if yes) | Ever vaccinated (1= if yes) |  |
|  | (1) | (2) | (3) | (4) | (5) | (6) | (7) | (8) | (9) |
| Treat × After | -0.049<br>(0.442) | 0.008<br>(0.318) | -0.033<br>(0.721) | -0.003<br>(0.064) | -0.022<br>(0.049) | 0.024<br>(0.068) | -0.014<br>(0.279) | -0.053<br>(0.066) | -0.025<br>(0.368) |
| Observations | 1,449 | 1,441 | 1,273 | 830 | 836 | 1,448 | 451 | 1,448 | 450 |
| State FE | Yes | Yes | Yes | Yes | Yes | Yes | Yes | Yes | Yes |
| Year FE | Yes | Yes | Yes | Yes | Yes | Yes | Yes | Yes | Yes |
| State-Year FE | Yes | Yes | Yes | Yes | Yes | Yes | Yes | Yes | Yes |
| Survey year FE | Yes | Yes | Yes | Yes | Yes | Yes | Yes | Yes | Yes |
| Controls | Yes | Yes | Yes | Yes | Yes | Yes | Yes | Yes | Yes |

### C. Heterogeneity

Next, we examine how the HSA affects child health among later-born children and daughters. Specifically, we test whether daughters or later-born children exhibit more significant health improvements. The HSA effects on human capital could differ by additional factors, such as gender and birth order. For example, the possible preference among parents for having a healthy eldest son could play a role. The issue pertains to Hindu families because of their patrilocal and patrilineal kinship systems. In addition, Hindu religious texts emphasize rituals that only a male heir can perform (Arnold, Choe, and Roy 1998).

To test for these effects, we re-estimate (1) by adding interaction terms for two demographic factors, gender and birth order:

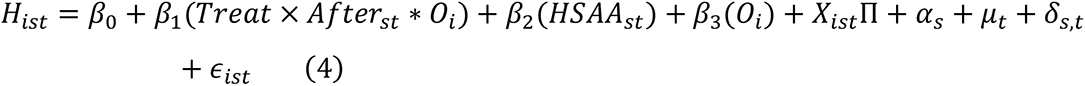

*O*_*i*_ denotes several demographic characteristics, such as birth order and gender. Specifically, we test whether the coefficient on the primary interaction is positive and statistically significantly different from zero.

We report the results in Tables 6 through 8. Table 6 reports the results from regressions estimated at the child level, and we examine the impact on the same set of outcomes as in Table 3. We focus on the coefficient of the interaction *Treat* × *After* × *Girl*. Columns 1, 2, and 9 report the estimates for the HFA z-score, WFA z-score, and the pool inputs index, respectively. We see a positive coefficient on *Treat* × *After* × *Girl*, but the coefficient is significant only for the WFA z-score outcome. Therefore, we can infer that the HSA has a positive effect on the daughter’s weight, but we cannot make the same claim for the HFA z-score and the parental inputs, as these two outcomes are not statistically significant.

**Table 6:**
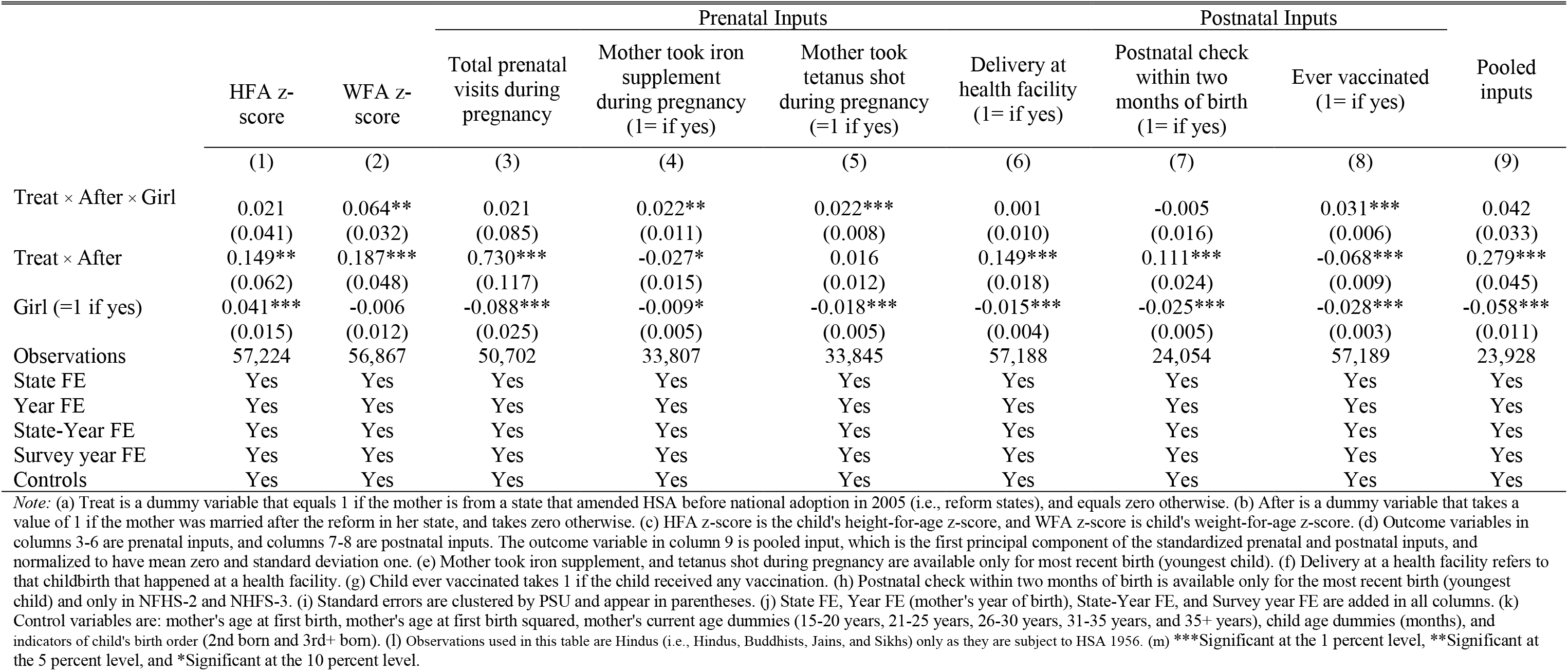
Heterogeneous Treatment Effect of HSAA on Child Health by Child’s Sex

**Table 7:** Heterogeneous Effect of HSAA on Child Health by Birth Order

|  | HFA z-score | WFA z-score | Prenatal Inputs |  |  | Postnatal Inputs |  |  | Pooled inputs |
| --- | --- | --- | --- | --- | --- | --- | --- | --- | --- |
|  |  |  | Total prenatal visits during pregnancy | Mother took iron supplement during pregnancy (1= if yes) | Mother took tetanus shot during pregnancy (=1 if yes) | Delivery at health facility (1= if yes) | Postnatal check within two months of birth (1= if yes) | Ever vaccinated (1= if yes) |  |
|  | (1) | (2) | (3) | (4) | (5) | (6) | (7) | (8) | (9) |
| Treat × After × 2nd Child | -0.021<br>(0.047) | 0.038<br>(0.035) | -0.231**<br>(0.094) | -0.012<br>(0.012) | 0.007<br>(0.009) | 0.030**<br>(0.012) | -0.040*<br>(0.021) | 0.009<br>(0.007) | -0.096**<br>(0.038) |
| Treat × After × 3rd+ Child | 0.003<br>(0.067) | 0.046<br>(0.050) | -0.293**<br>(0.132) | 0.027<br>(0.018) | 0.035***<br>(0.013) | 0.050***<br>(0.018) | -0.074***<br>(0.028) | 0.034***<br>(0.010) | -0.186***<br>(0.055) |
| Treat × After | 0.191***<br>(0.071) | 0.252***<br>(0.054) | 0.955***<br>(0.132) | -0.015<br>(0.018) | 0.015<br>(0.015) | 0.151***<br>(0.020) | 0.164***<br>(0.027) | -0.075***<br>(0.011) | 0.420***<br>(0.051) |
| 2nd Child (=1 if yes) | -0.246***<br>(0.021) | -0.227***<br>(0.017) | -0.898***<br>(0.041) | -0.066***<br>(0.008) | -0.047***<br>(0.006) | -0.186***<br>(0.005) | -0.080***<br>(0.008) | -0.037***<br>(0.004) | -0.300***<br>(0.018) |
| 3rd+ Child (=1 if yes) | -0.636***<br>(0.026) | -0.549***<br>(0.020) | -2.317***<br>(0.052) | -0.195***<br>(0.009) | -0.158***<br>(0.007) | -0.398***<br>(0.007) | -0.157***<br>(0.010) | -0.096***<br>(0.005) | -0.676***<br>(0.022) |
| Observations | 56,832 | 56,477 | 50,338 | 33,444 | 33,482 | 56,796 | 23,758 | 56,797 | 23,633 |
| State FE | Yes | Yes | Yes | Yes | Yes | Yes | Yes | Yes | Yes |
| Year FE | Yes | Yes | Yes | Yes | Yes | Yes | Yes | Yes | Yes |
| State-Year FE | Yes | Yes | Yes | Yes | Yes | Yes | Yes | Yes | Yes |
| Survey year FE | Yes | Yes | Yes | Yes | Yes | Yes | Yes | Yes | Yes |
| Controls | Yes | Yes | Yes | Yes | Yes | Yes | Yes | Yes | Yes |

**Table 8:** Heterogeneous Effect of HSAA on Child Health by Birth Order and Sex of the Children

|  | HFA z-score | WFA z-score | Prenatal Inputs |  |  | Postnatal Inputs |  |  | Pooled inputs |
| --- | --- | --- | --- | --- | --- | --- | --- | --- | --- |
|  |  |  | Total prenatal visits during pregnancy | Mother took iron supplement during pregnancy (1= if yes) | Mother took tetanus shot during pregnancy (=1 if yes) | Delivery at health facility (1= if yes) | Postnatal check within two months of birth (1= if yes) | Ever vaccinated (1= if yes) |  |
|  | (1) | (2) | (3) | (4) | (5) | (6) | (7) | (8) | (9) |
| Treat × After × 2nd Child × Girl | 0.083<br>(0.095) | -0.016<br>(0.076) | 0.001<br>(0.193) | 0.047*<br>(0.025) | 0.019<br>(0.018) | 0.028<br>(0.023) | 0.052<br>(0.039) | 0.022*<br>(0.012) | 0.086<br>(0.073) |
| Treat × After × 3rd+ Child × Girl | 0.191<br>(0.120) | 0.113<br>(0.094) | 0.063<br>(0.221) | 0.048<br>(0.032) | 0.019<br>(0.024) | 0.016<br>(0.032) | 0.016<br>(0.050) | 0.017<br>(0.017) | 0.040<br>(0.092) |
| Treat × After × 2nd Child | -0.062<br>(0.066) | 0.047<br>(0.050) | -0.232*<br>(0.134) | -0.034**<br>(0.017) | -0.001<br>(0.012) | 0.016<br>(0.016) | -0.065**<br>(0.027) | -0.000<br>(0.009) | -0.137***<br>(0.053) |
| Treat × After × 3rd+ Child | -0.087<br>(0.088) | -0.005<br>(0.068) | -0.324*<br>(0.165) | 0.005<br>(0.023) | 0.026<br>(0.017) | 0.043*<br>(0.024) | -0.082**<br>(0.038) | 0.026**<br>(0.012) | -0.205***<br>(0.072) |
| Treat × After × Girl | -0.060<br>(0.063) | 0.031<br>(0.050) | -0.029<br>(0.134) | -0.009<br>(0.017) | 0.002<br>(0.012) | -0.014<br>(0.015) | -0.025<br>(0.025) | 0.012<br>(0.008) | -0.015<br>(0.049) |
| 2nd Child × Girl | -0.118***<br>(0.041) | -0.058*<br>(0.032) | -0.074<br>(0.077) | -0.008<br>(0.015) | -0.009<br>(0.012) | -0.020*<br>(0.011) | -0.001<br>(0.015) | -0.009<br>(0.008) | -0.032<br>(0.033) |
| 3rd+ Child × Girl | -0.112***<br>(0.038) | -0.094***<br>(0.029) | -0.153**<br>(0.062) | -0.014<br>(0.013) | -0.033***<br>(0.011) | -0.015*<br>(0.009) | 0.006<br>(0.013) | -0.030***<br>(0.007) | -0.061**<br>(0.028) |
| Treat × After | 0.220***<br>(0.078) | 0.237***<br>(0.059) | 0.969***<br>(0.148) | -0.011<br>(0.019) | 0.014<br>(0.016) | 0.158***<br>(0.021) | 0.176***<br>(0.030) | -0.081***<br>(0.012) | 0.426***<br>(0.058) |
| 2nd Child (=1 if yes) | -0.189***<br>(0.030) | -0.199***<br>(0.023) | -0.862***<br>(0.055) | -0.062***<br>(0.010) | -0.043***<br>(0.008) | -0.176***<br>(0.008) | -0.080***<br>(0.011) | -0.032***<br>(0.005) | -0.284***<br>(0.024) |
| 3rd+ Child (=1 if yes) | -0.581***<br>(0.031) | -0.504***<br>(0.025) | -2.244***<br>(0.059) | -0.188***<br>(0.011) | -0.143***<br>(0.009) | -0.391***<br>(0.009) | -0.160***<br>(0.012) | -0.081***<br>(0.006) | -0.646***<br>(0.026) |
| Girl (=1 if yes) | 0.122***<br>(0.029) | 0.051**<br>(0.023) | -0.001<br>(0.055) | 0.001<br>(0.011) | 0.001<br>(0.008) | -0.003<br>(0.008) | -0.026**<br>(0.011) | -0.012**<br>(0.006) | -0.019<br>(0.024) |
| Observations | 56,832 | 56,477 | 50,338 | 33,444 | 33,482 | 56,796 | 23,758 | 56,797 | 23,633 |
| State FE | Yes | Yes | Yes | Yes | Yes | Yes | Yes | Yes | Yes |
| Year FE | Yes | Yes | Yes | Yes | Yes | Yes | Yes | Yes | Yes |
| State-Year FE | Yes | Yes | Yes | Yes | Yes | Yes | Yes | Yes | Yes |
| Survey year FE | Yes | Yes | Yes | Yes | Yes | Yes | Yes | Yes | Yes |
| Controls | Yes | Yes | Yes | Yes | Yes | Yes | Yes | Yes | Yes |

In Table 7, we report the estimations examining the effect of the HSA by birth order. The coefficient, Treat × After × 2nd Child and *Treat* × *After* × 3*rd* + *Child*, display the HSA effects on height (Columns 1), weight (Column 2), and parental investments (Column 9). We see a positive coefficient on *Treat* × *After* × *Girl*, but the coefficient is significant only for the WFA z-score outcome. For the health and weight outcomes, the coefficients on the interaction terms are not statistically significant. Strikingly, the signs of the coefficients for the parental investments associated with *Treat* × *After* × 2*nd Child* and *Treat* × *After* × 3*rd* + *Child* are not only negative but also statistically significant at the 5-percent level. Thus, the HSA has no positive effect for higher-born children and may have an adverse effect, via the parental allocations, on higher birth order children.

To test for both gender and birth order effects, we estimate (4) interacting the treatment with higher birth order and gender dummy variables. The coefficient of *Treat* × *After* × 2*nd Child* × *Girl* captures the policy effect among second-born daughters. The coefficient of *Treat* × *After* × 3*rd* + *Child* × *Girl* captures the policy effect among second-born daughters. Again, for none of the outcomes, the interactions are statistically significant, implying that the HSA reform did not necessarily benefit daughters of higher birth order.^15^

### D. Mechanisms

Although we have presented considerable evidence on how the HSA improves child health, we have not yet addressed the channels via which the HSAA likely improved health outcomes.

Several potential mechanisms can rationalize our findings. Modeling the household decisions for human capital investments is a challenging task. At the core of various explanations, which could explain our findings, is the non-unitary model of household bargaining and how changes in household bargaining influence subsequent parental investments into the human capital for children. A large body of evidence from developing countries supports the idea that households are not unitary entities and that bargaining power is crucial (Duflo, 2001; Qian, 2008). If we assume two parents in a non-unitary household model, the HSAA inheritance law changes positively influence the female bargaining power within the household. The HSAA allowed women to inherit property. It also increased their unearned income and raised their bargaining power (Heath and Tan, 2020).

The HSAA could lead to several channels conceptually consistent with an increase in child health. We first discuss each of these primary channels, and then we turn to the data to examine the empirical support for them. Using the variation due to the HSA implementation across states, we examine how the Act influenced each of these potential channels at the household level.^16^ We first examine the effect of the HSAA on each of the proposed channels. We estimate:

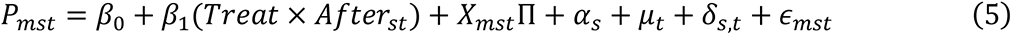

*P*_*ist*_ is a proxy measure for each of the proposed channels; *m* denotes the individual index in state *s* who was married in year *t*. We define all other variables as in the previous specifications.

A first possibility, because of the HSAA, is fertility stopping behavior and delaying childbearing. Suppose the wife gains bargaining power (conceptualized by having higher autonomy over household resources) due to the HSAA. In that case, the HSAA policy will likely reduce childbearing and delay childbearing as empirical evidence from India points to females having a lower preference for children and preference for bearing later. ((Bhattacharya, 2006).

The second channel relates to an increase of unearned income by women. The HSAA raises the woman’s level of unearned income due to enhanced access to the land inheritance. In a noncooperative household bargaining framework, this could translate to better control over the woman’s income and thereby better gains for the female in the household from her working. Heath and Tan (2020) show empirical evidence that the HSAA increase female labor supply. This increase in the wife’s labor supply did not come at the expense of the husband’s labor supply, allowing for a higher household income resulting from the HSAA. Additional household income may influence height through increasing calorie intake or the quality of the diet. Even if increased food intake does not directly come from the increase of total household income, reducing household size via reductions in childbearing (noted earlier) can lead to a higher nutritional intake per person.

Third, the disease environment in early childhood can play a critical role in influencing child health. Child height is a function of net nutrition, and net nutrition is the difference between food intake and the losses to activities and disease. In India, especially in rural parts, there is a high burden of diarrheal disease, fevers, or respiratory infections. These can impose a nutritive tax on one’s nutritional status in early childhood and subsequently on child height. Furthermore, there is clear evidence that inadequate prenatal nutrition, which is quite prevalent in India, causes low birth weight and worsens child health. Finally, infections during early childhood infections may have consequences for child height because they can sap energy required for physical growth. Both respiratory and gastrointestinal infections can impact height (Victora, 1990).

Finally, higher female bargaining power within the household can improve child outcomes. Existing research shows that an increase in women’s income or bargaining power within the household benefits children more than increases in men’s income (Thomas, 1990, Lundberg et al., 1997, Attanasio and Lechene, 2002). Households in which women’s income share is higher spend a more significant fraction of their income on children’s clothing and food, and children in these households are better fed. There are potentially two hypotheses put forth that could explain this phenomenon. The first relates to women and men having different preferences (the so-called preference hypothesis), with women being more concerned about their children and men’s well-being. This issue, combined with the assumption that women’s preferences matter more if their income share within the family goes up, can rationalize higher spending patterns on children’s needs, such as their health and education. According to this *preference hypothesis*, when giving transfers to women versus men, the trade-off is between additional spending on children versus private consumption of men. There is a second possibility called the *specialization hypothesis*. According to this hypothesis, differential spending can be explained by specialization by the household members. According to this framework, women and men specialize in different tasks based on their comparative advantage. For example, if women have lower wages than men, they specialize in time-intensive tasks, including childcare and food preparation. On the other hand, men will take charge of tasks that require money but little time, such as saving and investing.

### E. Supporting Evidence

Data limitations prevent us from taking on one particular channel against the others. Considering the assumptions required make a formal mediation analysis highly challenging in the context of our question. In what follows, we attempt to shed more light on the plausibility of some of the hypothesized channels using the available NFHS data. Table 9 reports the results from specification (5).

**Table 9:** Mechanisms of the Effect of HSAA on Child Health

|  | Fertility Choices |  | Household Wealth |  | Disease Environment | Female Bargaining |  |  |  |
| --- | --- | --- | --- | --- | --- | --- | --- | --- | --- |
|  | Total Children Born | Mother's Age at First Marriage | Electricity Access (=1 if yes) | Piped Water at Home (=1 if yes) | Childhood Disease Environment Index | Say About Large Purchase (=1 if yes) | Say About Own Health (=1 if yes) | Going to the Market Alone (=1 if yes) | Bargaining Index |
|  | (1) | (2) | (3) | (4) | (5) | (6) | (7) | (8) | (9) |
| Treat × After | -1.618***<br>(0.045) | 5.732***<br>(0.103) | 0.136***<br>(0.015) | 0.145***<br>(0.021) | -0.057<br>(0.038) | 0.103***<br>(0.020) | 0.107***<br>(0.018) | 0.077***<br>(0.019) | 0.267***<br>(0.037) |
| Observations | 46,495 | 46,495 | 45,174 | 46,495 | 46,422 | 32,490 | 32,491 | 32,709 | 32,490 |
| State FE | Yes | Yes | Yes | Yes | Yes | Yes | Yes | Yes | Yes |
| Year FE | Yes | Yes | Yes | Yes | Yes | Yes | Yes | Yes | Yes |
| State-Year FE | Yes | Yes | Yes | Yes | Yes | Yes | Yes | Yes | Yes |
| Survey year FE | Yes | Yes | Yes | Yes | Yes | Yes | Yes | Yes | Yes |
| Controls | Yes | Yes | Yes | Yes | Yes | Yes | Yes | Yes | Yes |

#### Childbearing

In Columns (1) and (2), we explore the childbearing channel with two outcomes: the total number of children ever born and the age at first marriage. The results reported in Table 9 show evidence consistent with the idea that among Hindu women living in HSAA-implementing states, the total number of children born decreased (in Column 1). Furthermore, there is evidence of delay in marriage (Column 2) and the first child’s timing (not reported). Overall, this fertility decline leads to reduced household size via reductions in childbearing, which are likely to result in higher nutritional intake per person.

#### Household Wealth

The IDHS does not collect income measures or any specific proxies of earnings by a household member. Therefore, we use two survey questions to proxy household wealth to capture the potential income effects. We rely on survey questions, whether the household has electricity and whether the household has piped water to proxy the change in household wealth in HSAA-implementing areas. Columns (3) and (4) in Table 9 report the results. The results show that both coefficients are positive and statistically significant, consistent with a positive income effect.

#### Disease Environment

In Column (5) of Table, we explore the possibility that the HSAA directly affects the disease environment in households. We show the results on a composite index based on the following variables: whether the child had a fever in the last two weeks, the incidence of cough in the last two weeks, and the incidence of diarrhea in the last two weeks. The coefficient displayed in column (5) is not statistically significant. Therefore, empirical evidence supports that the positive height impacts operate through a disease environment channel.

#### Female Bargaining Power

We explore the women’s bargaining channel using various empowerment measures available in the NFHS. Specifically, we use three decision measures: whether the woman has a say about large purchases (Column 6), whether she has a say about her own healthcare choices (Column 7), and whether she goes to the market alone (Column 8). We combine these measures using a composite female bargaining measure (Column 9). The coefficients associated with the female bargaining proxies are positive and statistically significant, implying that the female bargaining power increased due to the HSAA.

The above explanations are not necessarily mutually exclusive. Given the robust effects on the bargaining channel and the fact that higher wealth, through unearned income for the women due to the HSAAs, is likely to boost further female autonomy, we specifically examine the effects of the bargaining power channel on child health. We do so by augmenting the previous specification, and we use the implementation of the HSAA as an instrument for female bargaining power using the composite index.^17^ We estimate the following equation:

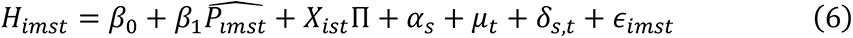

We define the variables as before. The HSAA instruments 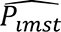, the female bargaining power.

Table 10 reports the results based on this approach. In columns (1) and (2), we show the impact on the HFA and WFA measures. In columns (3) through (6), we show the effect on prenatal inputs; columns (7) and (8) report the effects on postnatal inputs. Finally, column (9) reports the impacts on the composite index. The coefficients in columns (1), (2), and (9) are positive and statistically significant. Therefore, the results support the claim that higher female bargaining power at the household level improved child health in HSAA-implementing areas. We also examine whether the inclusion of other variables, particularly wealth proxies, knocks out the strong effect of the female bargaining power on child health (reported in Table 10). We conduct an additional exercise in which we interact wealth proxies with the bargaining power index and examine whether the effect size associated with the bargaining power variables changes after the inclusion of additional variables. The effect size of the bargaining power variable remains stable after the inclusion of these interaction terms. Although this exercise cannot rule out the possibility that wealth exerts an independent effect on child health, it strongly suggests that the bargaining power channel likely plays a critical role.

**Table 10:** Effect of Women’s Bargaining Power on Children’s Health

|  | HFA z-score | WFA z-score | Prenatal Inputs |  |  | Postnatal Inputs |  |  | Pooled inputs |
| --- | --- | --- | --- | --- | --- | --- | --- | --- | --- |
|  |  |  | Total prenatal visits during pregnancy | Mother took iron supplement during pregnancy (1= if yes) | Mother took tetanus shot during pregnancy (=1 if yes) | Delivery at health facility (1= if yes) | Postnatal check within two months of birth (1= if yes) | Ever vaccinated (1= if yes) |  |
|  | (1) | (2) | (3) | (4) | (5) | (6) | (7) | (8) | (9) |
| Bargaining | 0.446*<br>(0.252) | 0.949***<br>(0.224) | 2.868***<br>(0.553) | -0.060<br>(0.060) | 0.087*<br>(0.051) | 0.556***<br>(0.102) | 0.394***<br>(0.090) | -0.042<br>(0.034) | 1.010***<br>(0.179) |
| Observations | 39,711 | 39,492 | 33,261 | 33,233 | 33,270 | 39,680 | 23,662 | 39,677 | 23,538 |
| State FE | Yes | Yes | Yes | Yes | Yes | Yes | Yes | Yes | Yes |
| Year FE | Yes | Yes | Yes | Yes | Yes | Yes | Yes | Yes | Yes |
| State-Year FE | Yes | Yes | Yes | Yes | Yes | Yes | Yes | Yes | Yes |
| Survey year FE | Yes | Yes | Yes | Yes | Yes | Yes | Yes | Yes | Yes |
| Controls | Yes | Yes | Yes | Yes | Yes | Yes | Yes | Yes | Yes |
| 1<sup>st</sup> stage F-stat | 45.78 | 45.82 | 48.36 | 46.01 | 47.06 | 45.59 | 49.66 | 45.70 | 50.80 |

We also further drill down on how improved female bargaining affects children by various demographic characteristics. We specifically examine the effect of improved bargaining power by birth order and by the gender of the child. Although the effect for children of higher birth order or daughters is ambiguous dependent on the maternal preferences for these two characteristics, there is evidence from other settings (Duflo, 2003) that more income in the hands of mothers tends to benefit daughters more than sons. We report results based on specification (6) where we interact 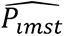 with additional covariates for birth-order and the gender of the child. Appendix A Table A2 and Table A3 report the results on child health using interactions for birth order and gender, respectively. The results show no evidence to support the claim that higher female bargaining power benefits either higher-order births or daughters more.

In sum, we cannot rule out the possibility that some of these channels in tandem improved child health. We also show robust empirical evidence that female bargaining power likely played a crucial role in improving child health because of the HSA.

## VI. Conclusion

Child height is a significant predictor of adult human capital and better economic outcomes. Nevertheless, India’s stunting rate stands at 31 percent, an astonishingly high prevalence rate even for developing countries. Moreover, because of the potential to transform women’s inheritance rights, some Indian states amended the male-favored Hindu Succession Act (HSA) by providing unmarried women an equal share in ancestral property. This paper studies the effect of the passage of inheritance law amendments (the HSAA), which conferred improved inheritance rights to unmarried women in India, on child height.

We exploit the staggered adoption of amendments to the HSA across different states in India since 1956. We find clear evidence that the passage of the HSAA improved, and substantially so, child health. To corroborate whether improved parental care drives these effects, we show that the HSA generated higher prenatal and postnatal investments in the areas that implemented the HSA amendments. We interpret these effects as mainly driven by two main channels: better female bargaining power and improved household wealth due to improvements in the wife’s unearned income. We show empirical support to claim that enhanced female autonomy at the household level can improve child health. This finding bolsters previous research showing that income in the hands of women (within the household) has a different impact on intra-household allocation than income in the hands of the men.

In developing countries such as India, improvements in human capital can offer a way to escape poverty. We explore several mechanisms, and we show that female empowerment can exert an important role in improving child health via improved inheritance rights. Moreover, our paper suggests that the enhanced inheritance rights for women may have far-reaching implications beyond their economic welfare, affecting human capital accumulation in the long run.

## Data Availability

Data will be made available upon request.

## Appendix A

**Table A1:**
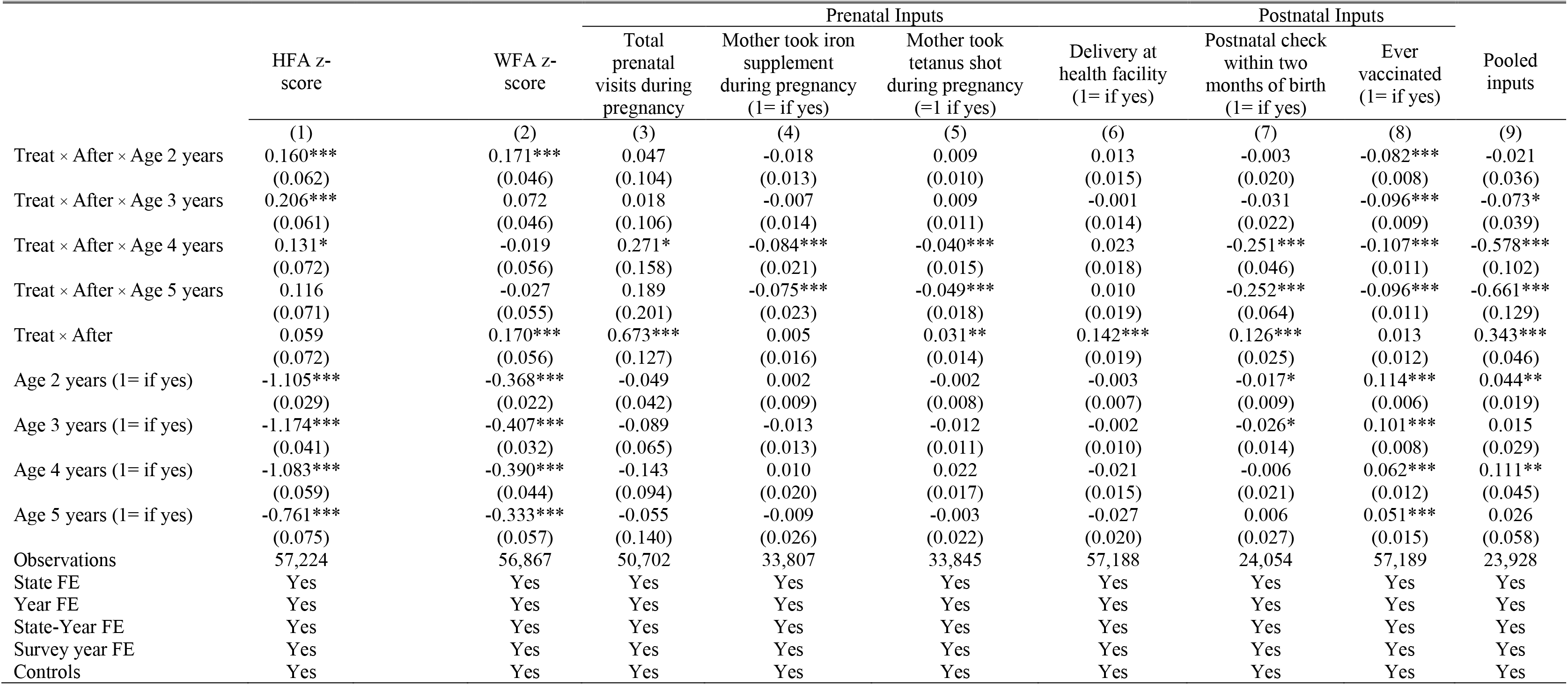
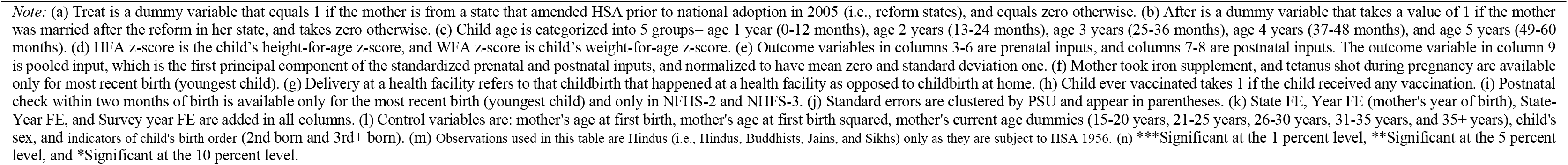
Heterogeneous Treatment Effect of HSAA on Child Health by Child’s Age

|  | HFA z-score | WFA z-score | Prenatal Inputs |  |  | Postnatal Inputs |  |  | Pooled inputs |
| --- | --- | --- | --- | --- | --- | --- | --- | --- | --- |
|  |  |  | Total prenatal visits during pregnancy | Mother took iron supplement during pregnancy (1= if yes) | Mother took tetanus shot during pregnancy (=1 if yes) | Delivery at health facility (1= if yes) | Postnatal check within two months of birth (1= if yes) | Ever vaccinated (1= if yes) |  |
|  | (1) | (2) | (3) | (4) | (5) | (6) | (7) | (8) | (9) |
| Treat × After × Age 2 years | 0.160***<br>(0.062) | 0.171***<br>(0.046) | 0.047<br>(0.104) | -0.018<br>(0.013) | 0.009<br>(0.010) | 0.013<br>(0.015) | -0.003<br>(0.020) | -0.082***<br>(0.008) | -0.021<br>(0.036) |
| Treat × After × Age 3 years | 0.206***<br>(0.061) | 0.072<br>(0.046) | 0.018<br>(0.106) | -0.007<br>(0.014) | 0.009<br>(0.011) | -0.001<br>(0.014) | -0.031<br>(0.022) | -0.096***<br>(0.009) | -0.073*<br>(0.039) |
| Treat × After × Age 4 years | 0.131*<br>(0.072) | -0.019<br>(0.056) | 0.271*<br>(0.158) | -0.084***<br>(0.021) | -0.040***<br>(0.015) | 0.023<br>(0.018) | -0.251***<br>(0.046) | -0.107***<br>(0.011) | -0.578***<br>(0.102) |
| Treat × After × Age 5 years | 0.116<br>(0.071) | -0.027<br>(0.055) | 0.189<br>(0.201) | -0.075***<br>(0.023) | -0.049***<br>(0.018) | 0.010<br>(0.019) | -0.252***<br>(0.064) | -0.096***<br>(0.011) | -0.661***<br>(0.129) |
| Treat × After | 0.059<br>(0.072) | 0.170***<br>(0.056) | 0.673***<br>(0.127) | 0.005<br>(0.016) | 0.031**<br>(0.014) | 0.142***<br>(0.019) | 0.126***<br>(0.025) | 0.013<br>(0.012) | 0.343***<br>(0.046) |
| Age 2 years (1= if yes) | -1.105***<br>(0.029) | -0.368***<br>(0.022) | -0.049<br>(0.042) | 0.002<br>(0.009) | -0.002<br>(0.008) | -0.003<br>(0.007) | -0.017*<br>(0.009) | 0.114***<br>(0.006) | 0.044**<br>(0.019) |
| Age 3 years (1= if yes) | -1.174***<br>(0.041) | -0.407***<br>(0.032) | -0.089<br>(0.065) | -0.013<br>(0.013) | -0.012<br>(0.011) | -0.002<br>(0.010) | -0.026*<br>(0.014) | 0.101***<br>(0.008) | 0.015<br>(0.029) |
| Age 4 years (1= if yes) | -1.083***<br>(0.059) | -0.390***<br>(0.044) | -0.143<br>(0.094) | 0.010<br>(0.020) | 0.022<br>(0.017) | -0.021<br>(0.015) | -0.006<br>(0.021) | 0.062***<br>(0.012) | 0.111**<br>(0.045) |
| Age 5 years (1= if yes) | -0.761***<br>(0.075) | -0.333***<br>(0.057) | -0.055<br>(0.140) | -0.009<br>(0.026) | -0.003<br>(0.022) | -0.027<br>(0.020) | 0.006<br>(0.027) | 0.051***<br>(0.015) | 0.026<br>(0.058) |
| Observations | 57,224 | 56,867 | 50,702 | 33,807 | 33,845 | 57,188 | 24,054 | 57,189 | 23,928 |
| State FE | Yes | Yes | Yes | Yes | Yes | Yes | Yes | Yes | Yes |
| Year FE | Yes | Yes | Yes | Yes | Yes | Yes | Yes | Yes | Yes |
| State-Year FE | Yes | Yes | Yes | Yes | Yes | Yes | Yes | Yes | Yes |
| Survey year FE | Yes | Yes | Yes | Yes | Yes | Yes | Yes | Yes | Yes |
| Controls | Yes | Yes | Yes | Yes | Yes | Yes | Yes | Yes | Yes |

**Table A2:** Effect of Women’s Bargaining Power on Child Health

|  | HFA z-score | WFA z-score | Prenatal Inputs |  |  | Postnatal Inputs |  |  | Pooled inputs |
| --- | --- | --- | --- | --- | --- | --- | --- | --- | --- |
|  |  |  | Total prenatal visits during pregnancy | Mother took iron supplement during pregnancy (1= if yes) | Mother took tetanus shot during pregnancy (=1 if yes) | Delivery at health facility (1= if yes) | Postnatal check within two months of birth (1= if yes) | Ever vaccinated (1= if yes) |  |
|  | (1) | (2) | (3) | (4) | (5) | (6) | (7) | (8) | (9) |
| Panel A: Hindu |  |  |  |  |  |  |  |  |  |
| Bargaining × 2nd Child | 0.059<br>(0.205) | 0.231<br>(0.184) | -0.742<br>(0.535) | -0.047<br>(0.052) | 0.034<br>(0.036) | 0.208***<br>(0.075) | -0.103<br>(0.176) | 0.005<br>(0.028) | -0.289<br>(0.368) |
| Bargaining × 3rd+ Child | 0.181<br>(0.290) | 0.168<br>(0.256) | -0.887<br>(0.659) | 0.101<br>(0.067) | 0.108**<br>(0.052) | 0.257**<br>(0.120) | -0.473***<br>(0.142) | 0.044<br>(0.039) | -1.195***<br>(0.311) |
| Bargaining | 0.395<br>(0.284) | 0.821***<br>(0.250) | 3.367***<br>(0.652) | -0.054<br>(0.069) | 0.053<br>(0.058) | 0.425***<br>(0.116) | 0.566***<br>(0.125) | -0.050<br>(0.039) | 1.455***<br>(0.265) |
| 2nd Child (=1 if yes) | -0.259***<br>(0.024) | -0.200***<br>(0.021) | -0.952***<br>(0.063) | -0.069***<br>(0.007) | -0.042***<br>(0.005) | -0.177***<br>(0.009) | -0.097***<br>(0.030) | -0.026***<br>(0.004) | -0.350***<br>(0.061) |
| 3rd+ Child (=1 if yes) | -0.641***<br>(0.041) | -0.476***<br>(0.038) | -2.309***<br>(0.103) | -0.196***<br>(0.012) | -0.139***<br>(0.010) | -0.364***<br>(0.018) | -0.207***<br>(0.025) | -0.071***<br>(0.006) | -0.804***<br>(0.053) |
| Observations | 39,711 | 39,492 | 33,261 | 33,233 | 33,270 | 39,680 | 23,662 | 39,677 | 23,538 |
| State FE | Yes | Yes | Yes | Yes | Yes | Yes | Yes | Yes | Yes |
| Year FE | Yes | Yes | Yes | Yes | Yes | Yes | Yes | Yes | Yes |
| State-Year FE | Yes | Yes | Yes | Yes | Yes | Yes | Yes | Yes | Yes |
| Survey year FE | Yes | Yes | Yes | Yes | Yes | Yes | Yes | Yes | Yes |
| Controls | Yes | Yes | Yes | Yes | Yes | Yes | Yes | Yes | Yes |
| 1<sup>st</sup> stage F-stat | 30.10 | 29.23 | 29.98 | 29.86 | 29.92 | 30.05 | 20.47 | 30.15 | 20.33 |

**Table A3:** Effect of Women’s Bargaining Power on Child Health

|  | HFA z-score | WFA z-score | Prenatal inputs |  |  | Postnatal Inputs |  |  | Pooled inputs |
| --- | --- | --- | --- | --- | --- | --- | --- | --- | --- |
|  |  |  | Total prenatal visits during pregnancy | Mother took iron supplement during pregnancy (1= if yes) | Mother took tetanus shot during pregnancy (=1 if yes) | Delivery at health facility (1= if yes) | Postnatal check within two months of birth (1= if yes) | Ever vaccinate d (1= if yes) |  |
|  | (1) | (2) | (3) | (4) | (5) | (6) | (7) | (8) | (9) |
| Panel A: Hindu |  |  |  |  |  |  |  |  |  |
| Bargaining × 2nd Child × Girl | 0.252<br>(0.404) | -0.012<br>(0.380) | 0.325<br>(1.045) | 0.187*<br>(0.102) | 0.082<br>(0.072) | 0.240<br>(0.163) | 0.228<br>(0.341) | 0.077<br>(0.053) | 0.321<br>(0.729) |
| Bargaining × 3rd+ Child × Girl | 0.524<br>(0.464) | 0.263<br>(0.414) | -0.293<br>(1.064) | 0.155<br>(0.114) | 0.023<br>(0.086) | -0.068<br>(0.189) | 0.033<br>(0.253) | 0.067<br>(0.064) | 0.039<br>(0.541) |
| Bargaining × 2nd Child | -0.052<br>(0.271) | 0.240<br>(0.234) | -0.888<br>(0.679) | -0.127*<br>(0.070) | -0.002<br>(0.046) | 0.101<br>(0.097) | -0.223<br>(0.225) | -0.028<br>(0.037) | -0.497<br>(0.459) |
| Bargaining × 3rd+ Child | -0.063<br>(0.372) | 0.043<br>(0.314) | -0.737<br>(0.847) | 0.034<br>(0.088) | 0.100<br>(0.065) | 0.296*<br>(0.166) | -0.477**<br>(0.191) | 0.014<br>(0.050) | -1.200***<br>(0.389) |
| Bargaining × Girl | -0.034<br>(0.253) | 0.283<br>(0.225) | 0.106<br>(0.757) | -0.043<br>(0.067) | 0.016<br>(0.049) | -0.035<br>(0.086) | -0.035<br>(0.176) | 0.003<br>(0.032) | 0.171<br>(0.425) |
| 2nd Child × Girl | -0.073<br>(0.046) | -0.050<br>(0.042) | -0.107<br>(0.130) | -0.001<br>(0.013) | -0.008<br>(0.010) | -0.013<br>(0.018) | 0.043<br>(0.053) | -0.004<br>(0.007) | 0.021<br>(0.117) |
| 3rd+ Child × Girl | -0.049<br>(0.046) | -0.109***<br>(0.041) | -0.190<br>(0.125) | 0.002<br>(0.013) | -0.028***<br>(0.010) | -0.008<br>(0.018) | 0.011<br>(0.045) | -0.022***<br>(0.007) | -0.070<br>(0.097) |
| Bargaining | 0.349<br>(0.319) | 0.611**<br>(0.275) | 3.326***<br>(0.726) | -0.052<br>(0.076) | 0.038<br>(0.062) | 0.452***<br>(0.127) | 0.570***<br>(0.143) | -0.061<br>(0.043) | 1.331***<br>(0.297) |
| 2nd Child (=1 if yes) | -0.223***<br>(0.033) | -0.174***<br>(0.029) | -0.896***<br>(0.089) | -0.067***<br>(0.010) | -0.037***<br>(0.007) | -0.168***<br>(0.013) | -0.119***<br>(0.034) | -0.023***<br>(0.005) | -0.364***<br>(0.069) |
| 3rd+ Child (=1 if yes) | -0.621***<br>(0.048) | -0.430***<br>(0.042) | -2.220***<br>(0.124) | -0.198***<br>(0.014) | -0.127***<br>(0.011) | -0.359***<br>(0.022) | -0.213***<br>(0.033) | -0.061***<br>(0.007) | -0.773***<br>(0.067) |
| Girl (=1 if yes) | 0.065**<br>(0.032) | 0.042<br>(0.028) | -0.020<br>(0.102) | -0.004<br>(0.009) | 0.001<br>(0.006) | -0.011<br>(0.011) | -0.037<br>(0.036) | -0.002<br>(0.004) | 0.009<br>(0.085) |
| Observations | 39,711 | 39,492 | 33,261 | 33,233 | 33,270 | 39,680 | 23,662 | 39,677 | 23,538 |
| State FE | Yes | Yes | Yes | Yes | Yes | Yes | Yes | Yes | Yes |
| Year FE | Yes | Yes | Yes | Yes | Yes | Yes | Yes | Yes | Yes |
| State-Year FE | Yes | Yes | Yes | Yes | Yes | Yes | Yes | Yes | Yes |
| Survey year FE | Yes | Yes | Yes | Yes | Yes | Yes | Yes | Yes | Yes |
| Controls | Yes | Yes | Yes | Yes | Yes | Yes | Yes | Yes | Yes |
| 1<sup>st</sup> stage F-stat | 15.76 | 15.25 | 15.49 | 15.46 | 15.54 | 15.74 | 11.47 | 15.81 | 11.21 |

**Table A4:** Effect of Bargaining on Child Health

|  | HFA z-score |  |  | WFA z-score |  |  |
| --- | --- | --- | --- | --- | --- | --- |
|  | OLS | IV | Imperfect<br>IV (Upped<br>Bound) | OLS | IV | Imperfect<br>IV (Upped<br>Bound) |
|  | (1) | (2) | (3) | (4) | (5) | (6) |
| Bargaining | 0.048***<br>(0.010) | 0.446*<br>(0.252) | 0.053<br>[0.072] | 0.041***<br>(0.008) | 0.949***<br>(0.224) | 0.048<br>[0.061] |
| Observations | 40,101 | 39,711 | 39,763 | 39,880 | 39,492 | 39,546 |
| State FE | Yes | Yes | Yes | Yes | Yes | Yes |
| Year FE | Yes | Yes | Yes | Yes | Yes | Yes |
| State-Year FE | Yes | Yes | Yes | Yes | Yes | Yes |
| Survey year FE | Yes | Yes | Yes | Yes | Yes | Yes |
| Controls | Yes | Yes | Yes | Yes | Yes | Yes |

## Footnotes

† We thank Declan Levine, Matthew Bonci, Justin Sun, Ariunzaya Oktyabri, and Charlotte Williams for outstanding research assistance. We thank Julie Cullen, Subal Kumbhakar, Eric Edmonds, Lauren Bergquist, Rachel Heath, Agnitra Roy Choudhury, Paul Novosad, Angela de Oliveira, Lisa Kahn, Seema Jaychandran, Belinda Archibong, Sulagna Mookerjee, Livia Montana, Jessica Goldberg, S. Anukriti, Petra Moser, Susan Wolcott, Evan Riehl, Nusrat Jimi, Shiv Hastawala, and Solomon Polachek for constructive feedback and helpful comments.

1 Several studies examine the role of genetic factors (Myres et al., 2011; Rootsi et al., 2004), disease environment (Bozzoli, Deaton, Quintana-Domeque, 2009). Other studies focus on the role cultural gender preference and unequal resource allocations within a family based on perceived returns to investment (Rosenzweig and Schultz 1982; Behrman 1988; Oster 2009; Jayachandran and Pande 2017).

2 One caveat could be that parents can purposefully advance (those who want to avoid devolving property to daughters) or delay (those who are gender progressive) daughters’ marriage (Bose and Das, 2017). This sort of selection in the marriage timing is not necessariy a concern as the amendments were often implemented retrospectively. Similar to Bose and Das (2017), we account for the age at marriage to account for any potential bias.

3 The patrilineal system stimulates preference for son as sons tend to reside with parents, take on the responsibilities of the parents in their old age, work on the family land, and subsequently inherit it. In contrast, daughters marry some distance from their natal home and take with them family assets as dowry (Rosenzweig and Wolpin 1985). These is existing evidence that son-preference is stronger in families where the first-born child is not a son; these families tend to be larger as they keep growing until their desired gender mix of children is achieved (Clark, 2000).

4 However, both sibling composition in the household and resource constraints can play a critical role in resource distribution among children, particularly female children (Haan, Plug, and Rosero, 2014). Studies showed daughters in families without a son are breastfeed for a shorter period (Haan, Plug, and Rosero, 2014; Jayachandran and Kuziemko, 2011).

5 A daughter with no elder brother may benefit from a lack of sibling rivalry. Still, the fertility-stopping effect reduces resources received by the daughter as the family realizes that they need to try again for a son and start saving funds for the next child (an expected son). Jayachandran and Pande (2017) find that the fertility-stopping effect dominates, and a daughter with no brother receives relatively fewer postnatal health inputs than prenatal inputs.

6 Existing research documents the effects of the HSAA on the likelihood to inherit land (Deininger, 2013), labor supply and placements into high-paying jobs (Heath and Tan, 2020), and educational attainment (Roy, 2015; Bose and Das, 2017). There is also evidence of some negative consequences on domestic violence (wife beating), increased suicide rate (Anderson and Genicot, 2015), increased female child mortality (Rosenblum 2015), and higher sex-selective abortion (Bhalotra et al., 2017).

7 Joint-family is a legal term, not necessarily requiring sharing the same household.

8 Goyal et al. (2013) argues that the proportion of people who die without preparing a will is very high in India (around 65 percent, the share is even higher in rural areas).

9 The HSA applied to all states except for Jammu and Kashmir, a region that administered its version of the Act. Although the Act had special provisions for the matrilineal communities, tribal communities of the Northeastern states were excluded as they were matrilineal but ruled by local customs (Agarwal, 1994).

10 The reforms in Kerala were quite distinct from the other state-level reforms, as they abolished the system of joint property altogether (Roy, 2015; Bose and Das, 2017).

11 The amendments were often implemented retrospectively (Mookerjee, 2019). For example, Andhra Pradesh formally passed the amendment on May 1986 but it was retrospectively in effect from September 1985 onwards.

12 We exclude Union territories, West Bengal, Jammu and Kashmir, Kerala, and Northeastern states from our analysis. Union territories are politically and administratively different from rest of India; West Bengal and Assam practice the Deyabhaga system of property right, which allows girls to inherit various types of property. Jammu and Kashmir was not subject to the HSA. Kerala and the eight northeastern states are not part of the analysis because they are matrilineal kinship areas (Jayachandran and Pande 2017).

13 A z-score of 0 represents the median of the reference population, and a z-score of -2 indicates that the child is two standard deviations below the reference population mean.

14 A potential concern could be out-migration to different states from the state the woman was born. However, cultural and linguistic barriers impede cross-state migration in India; cross-state migration is negligible (Roy, 2015; Bose and Das, 2017).

15 In a separate extension exercise, we also examine whether the timing of the HSAA influenced child health differentially by child age. We report the results of this estimation in Appendix Table A1. There is suggestive evidence, based on the results reported, that the policy has a bigger effect size in earlier childhood years.

16 Heath and Tan (2019) show that the HSAA significantly improved female bargaining power.

17 Although we use the HSAA as an instrument for bargaining power, we conduct an additional robustness exercise. In this exercise, we adopt the approach proposed by Nevo & Rosen (2012), and we allow for the instrument to be correlated with the error term. However, we assume that the correlation is weaker than the correlation between the instrumental variable and the endogenous variable itself. Using the econometric technique, proposed by Nevo & Rosen (2012), we bound the parameter of interest. The purpose of this additional consistency check is to ascertain the bounds the parameter of interest in case the instrumental variable does not fulfill the classical exclusion restriction assumption. Appendix A, Table A4 reports the results. The reported bounds indicate that the results reported in Table 7 are robust to relaxing the exclusion restriction assumption.

